# Closed-loop auditory stimulation (CLAS) does not improve sleep or declarative memory in chronic insomnia

**DOI:** 10.1101/2025.03.04.25321710

**Authors:** Aurore A. Perrault, Ju Lynn Ong, Emma-Maria Phillips, Nathan E. Cross, Teck Boon Teo, Andrew R. Dicom, Nicholas I.Y.N. Chee, Amiya Patanaik, Michael W.L. Chee, Thien Thanh Dang Vu

## Abstract

**Objective:** Investigate whether auditory closed-loop stimulation (CLAS) applied during sleep could be beneficial for sleep and declarative memory in individuals with chronic insomnia.

**Methods:** We performed a randomized crossover sham-controlled study on 27 individuals with chronic insomnia to assess changes in sleep and declarative memory between a night with CLAS (i.e., 2-ON-OFF blocks auditory tones locked to slow wave up-states during NREM) and a SHAM night. We conducted assessments of memory (word paired-associate learning task) and sleep (morning questionnaire, polysomnographic recordings) during both nights.

**Results:** We found that applying CLAS in a population of individuals with chronic insomnia led to an acute increase in SO amplitude after auditory stimulation. However, we found no beneficial effect of a single night of CLAS on subjective and objective sleep or declarative overnight memory performance. There was an association between the increase in SO density during CLAS with fewer markers of sleep fragmentation (i.e., sleep fragmentation index, arousals), suggesting interindividual differences in response to CLAS in chronic insomnia.

**Conclusions:** CLAS stimulation applied during NREM sleep in individuals with chronic insomnia is feasible but did not show consistent effects on EEG markers of sleep regulation. A subgroup of individuals with insomnia may be more responsive to the impact of CLAS on sleep maintenance.

## BACKGROUND

The exposure to rhythmic auditory stimulation in phase with ongoing endogenous neural slow oscillations (<1Hz) during sleep, also called closed-loop auditory stimulation (CLAS), has been mostly used to manipulate brain oscillations that regulate sleep-dependent memory processes in good sleepers[1]. To date, multiple studies have shown that CLAS bestows general beneficial effects on memory performance[2,3]. Although most studies showed no change in sleep macro-architecture, CLAS promotes the occurrence of slow oscillations (SOs) and increases delta and spindle activity[2–6]. Precisely timed CLAS and the associated increase in SO amplitude may have a beneficial role in protecting sleep continuity in the face of noise[7]. As such, CLAS may constitute a promising method of promoting sleep maintenance and cognition in populations with cognitive impairment[8] or sleep difficulties[4,9]. Recent use of CLAS in clinical populations revealed the feasibility of CLAS in-lab and at-home[4,9–11] with some improvement in alertness and attention in short sleepers[4]. Yet there is very little data on the effects of CLAS on sleep and memory consolidation in chronic insomnia. Chronic insomnia is defined by complaints of difficulty falling asleep and/or maintaining sleep, occurring at least 3 times per week for more than 3 months, and is accompanied by daytime functioning complaints such as fatigue, mood disruption and cognitive complaints (e.g., concentration, memory)[12]. A recent pilot study showed an increase in slow oscillations but a reduction in spindle activity with no effect on sleep architecture and memory consolidation in a small sample of 7 individuals with chronic insomnia[9].

This study aimed to extend these recent findings to a larger sample, investigating whether CLAS could be beneficial for the sleep and declarative memory of individuals with chronic insomnia.

## PATIENTS AND METHODS

We performed a randomized crossover sham-controlled study on 27 individuals with chronic insomnia. Inclusion criteria were being over 21 years old, meeting the DSM-V criteria for chronic insomnia [13] and being a native english speaker. Exclusion criteria included the use of active pharmacological agents that could affect sleep, other sleep disorders, history of severe medical illness or neurological disorders, shift work, formal education <12 years, consume more than 2 units (200mg) of caffeine a day, consume more than 21 units of alcohol per week, travelling across >1 timezone in the past month and a Mini-Mental State Examination (MMSE) <27. The study was approved by the Institutional Review Board of the National University of Singapore (reference H-18-004), registered as a clinical trial (ISRCTN73971514), and all participants signed an informed consent form.

Participants underwent a habituation night followed by two experimental nights with polysomnographic (PSG) recording in a counterbalanced order: one with CLAS stimulation (STIM) and one without stimulation (SHAM). The CLAS method has been described elsewhere[3,5]. In brief, we used real-time automated sleep stage detection and up-state targeting of SOs during N2 and N3 on electrode F3:A2. During STIM night, auditory tones (50ms bursts of pink noise) locked to SO upstates were played in 2-ON-OFF blocks. Tone presentation was halted if an arousal occurred or if voltage thresholds (40uV) were not met. During the SHAM night, the SO up-state locations were detected but no tones were played.

Four subjects were removed from all analyses because their SO amplitudes were below threshold (40uV) to trigger/mark the stimulation. After each night, participants filled out a short survey on the quality of their sleep, sleep duration, number of awakenings and whether they perceived sounds during the night.

PSG recordings included 10-channel EEG sampled at 512 Hz, EOG and EMG (Brain Products, Germany). Sleep stages, artefacts and arousals were scored by 2 blind scorers (AAP, EMP) according to AASM guidelines[15] using the Wonambi python toolbox (https://wonambi-python.github.io) and analyses were conducted using the SEAPIPE python toolbox (https://github.com/nathanecross/seapipe).

To assess the evoked activity of CLAS, the EEG signal for both STIM and SHAM conditions was bandpass averaged across all ON blocks within a 4-s window (100s bins), time-locked to the first of the 2 stimuli ON block and a pre-stimulus onset time of 1-s.

The EEG spectrum power average (30-s of time resolution with artefacts excluded) was calculated with a 0.2 Hz resolution, by applying a Fast Fourier Transformation (FFT; 50 % overlapping, 5-s windows, Hanning filter) on F3:A2. Analyses in the current report focused on relative SO (0.25-1.25Hz) and delta (0.25-4 Hz) spectral power (band/total power) during N2 and N3 sleep epochs Spindle detection was based on the procedure of Ray and colleagues[16] on F3:A2 and C3:A2, while SOs were detected following the procedure of Ngo and colleagues[2] on frontal derivations (F3:A2, F4:A1).

Finally, to assess the impact of CLAS stimulation on overnight declarative memory performance, we used a word paired-associate learning task described previously[3]. Encoding and immediate recall were performed in the evening until an accuracy criterion of 60% was reached, while delayed recall was performed in the morning. Memory accuracy (correct minus wrong) and overnight memory accuracy (morning minus evening) were extracted from the last immediate test and the delayed recall test in the morning for each night.

Statistical analyses were conducted using RStudio 1.2.50 (RStudio, Inc., Boston, MA) and R custom scripts and functions (e.g., packages nparLD, rstatix, car). Normality of distribution was checked with Shapiro tests and the homogeneity of variance was tested with Levene tests. Effects were measured using repeated-measures analyses of variances (ANOVA) and paired t-tests or their non-parametric versions (paired Wilcox Test). All tests included a repeated measure factor Condition (SHAM, STIM) with Age as covariate. We computed effect sizes to indicate the degree of change in response to Condition using Hedges’s *g* (corrects for small sample size). Exploratory Pearsons’ correlations were conducted between changes in sleep and memory variables. The level of significance was set to p <.05.

## RESULTS

Twenty-three individuals with chronic insomnia were analyzed in this crossover design. Participants were middle-aged adults (mean: 33 ± 15 years old; 65.2% females) suffering from chronic insomnia with mild to moderate insomnia severity as measured by the insomnia severity index [14] (mean score 15.9 ± 1.8; **Figure 1A**).

**Figure 1.**
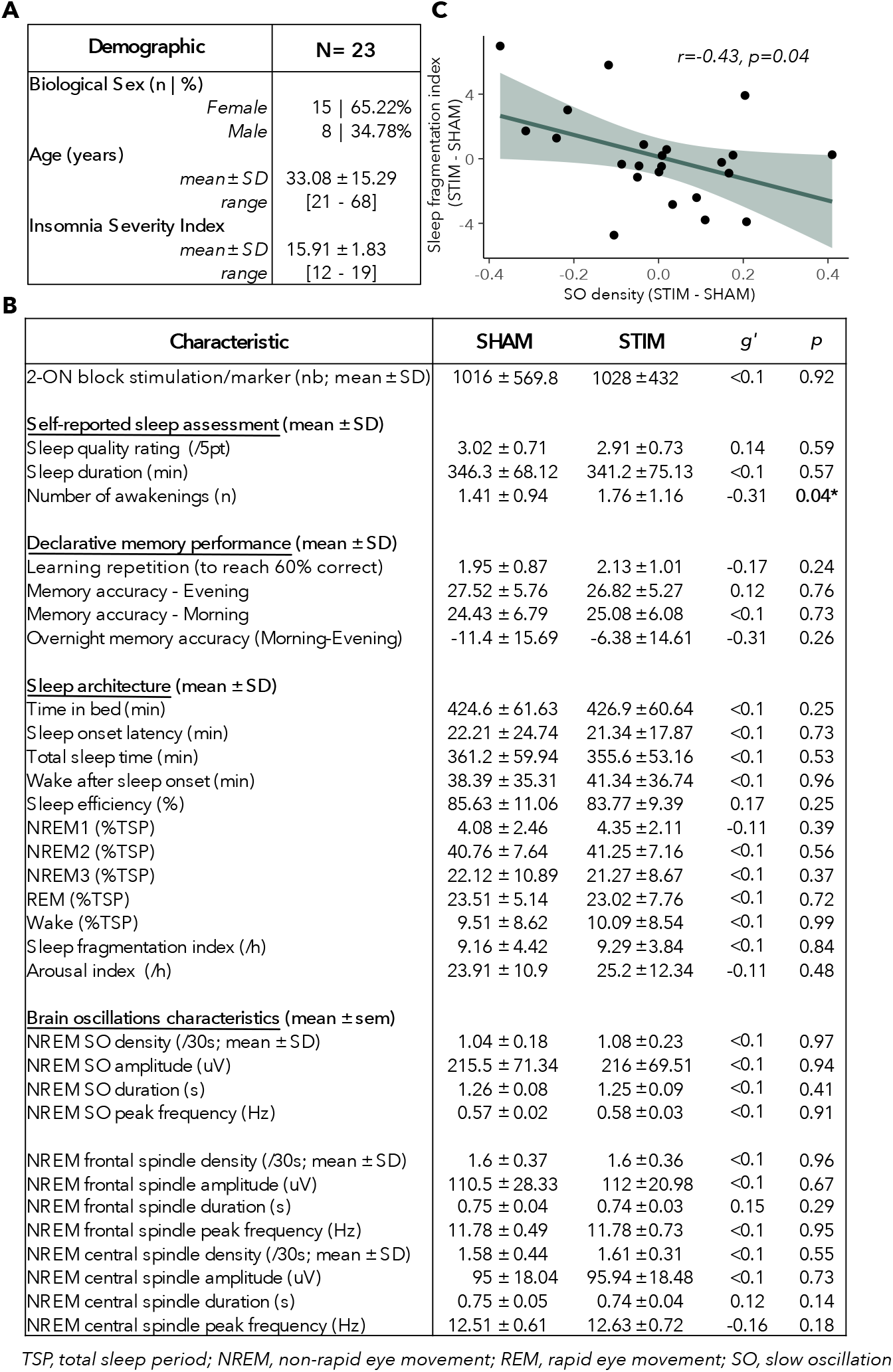
CLAS does not improve sleep or declarative memory in chronic insomnia. *A) Demographic measures* *B) Sleep and memory measures during STIM and SHAM night* *C) Scatter plot showing a significant correlation between the change (STIM minus SHAM) in SO density and sleep fragmentation index*

On average, participants received 1028 (± 432) auditory simulations (2-ON blocks) during NREM sleep in the STIM condition, compared with 1015 (± 569) marked as 2-blocks of SO up-states in the SHAM condition. While this difference was not significant at the group level (F(1,21) = 0.010; *P* =.92), we found that the degree of change in the number of 2-block markers/stimulations between SHAM and STIM ranged from −1013 to +632, suggesting a strong inter-individual difference in CLAS response. Moreover, we found a large effect of Age (F(1,21) = 31.15; *P* <.001) on the number of 2-blocks received/marked, where younger individuals received more 2-blocks stimulation than older adults (STIM: r=−0.75, p<.001; SHAM: r=−0.65, p=.002). However, there was no association between ISI severity and number of 2-blocks received/marked (all p>.05).

### CLAS DOES NOT IMPROVE SUBJECTIVE SLEEP QUALITY AND OVERNIGHT MEMORY PERFORMANCE IN INDIVIDUALS WITH INSOMNIA

When asked in the morning to rate their sleep quality, participants did not report having a better sleep quality after the STIM night compared to SHAM night (p=.59). However, they reported remembering more awakenings during the STIM night (+24.8%; p=.044, g’=−0.3). There was no effect of Condition on overnight memory accuracy (p=.26; **Figure 1B**).

### CLAS DOES NOT AFFECT SLEEP MACRO-AND MICRO-ARCHITECTURE

Investigation of sleep architecture using PSG recording (e.g., sleep efficiency, total wake and sleep time, sleep latency, percentage of time spent in each stage) did not reveal any effect of Condition (all p>.05, all g’<0.1). There was also no change in the markers of sleep fragmentation (i.e., sleep fragmentation index, arousal density; all p>.05, all g’<0.1; **Figure 1B**).

While stimulation elicited a greater acute increase in the amplitude of SO activity after ON1 in the STIM condition compared to SHAM (all bins p<.05 up to 1.2s post-ON1), we found that the proportion of spindles and SOs occurring after the 2-blocks stimulation/marking were not different between conditions (all p>.05). Moreover, whole night analyses revealed no effect of Condition on SO and delta relative power during REM and NREM sleep (combined N2+N3 and per stage; all p>.05). Finally, we found that CLAS stimulation did not alter the overall quantity or characteristics (i.e., density, frequency, amplitude, duration) of NREM frontal and central spindles and NREM SO events compared to SHAM night (all p>.05 and g’<0.2; **Figure 1B**).

Interestingly, we found a correlation between changes in SO density during NREM and changes in markers of sleep fragmentation. Indeed, individuals who exhibited an increase in NREM SO density with CLAS displayed less NREM arousal density (r=−0.44, p=0.03) and reduced sleep fragmentation indices [17] (r=−0.43, p=0.04; **Figure 1C**). However, this was not associated with better subjective sleep quality, memory performance or any other sleep measures (all p>.05).

## DISCUSSION

While the application of CLAS in a population of individuals with chronic insomnia is feasible (i.e., acute increase in SO amplitude after auditory stimulation), we found no beneficial effect of a single night of CLAS on subjective and objective sleep or declarative memory performance. Our results are congruent with a recent pilot study conducted by Dudysova and colleagues[9]. They did not find any change in sleep architecture or memory performance between a night of CLAS and a night of SHAM in a small sample of individuals with chronic insomnia. With more than half of their participants (10 out of 17) not receiving enough acoustic stimulations during sleep (<50 stimulations), the authors discussed the need to adapt CLAS methodology (e.g., amplitude threshold) on individuals with insomnia to account for morphological differences in sleep-related brain oscillations. While the low number of acoustic stimulations might have explained their lack of change, in the present study, 85% of our participants (23/27) received more than 300 acoustic stimulations during NREM, suggesting further that CLAS does not alter sleep quality in insomnia. However, similar to previous studies on healthy and clinical populations[4,10,18], we also observed a large heterogeneity in the number of acoustic stimulations received (mean 981 ± 399 during CLAS night). The inter-individual difference in the number of acoustic stimulations delivered was not associated with insomnia severity but rather with age, similar to a previous study in young and older healthy sleepers[18]. This is likely explained by a decrease in SO amplitude that is associated with aging[19].

While the absence of changes in sleep macro-architecture during CLAS has been reported in some studies[2,3,5,18], we also did not find any changes in NREM brain oscillations occurrence. The acute effect of CLAS led to an increase in SO amplitude but overall, we failed to replicate the increase in delta or sigma power often reported by CLAS studies[2,3,5,6] although not always observed[9,18]. Aligning with the current theory of sleep-dependent memory consolidation processes[20], the absence of change in NREM response patterns may also explain the lack of changes in declarative memory performance in our sample of chronic insomnia. It is possible that because healthy sleepers usually have better sleep at baseline than those with sleep difficulties, the beneficial effects on brain oscillations and memory consolidation in patients with insomnia might require more than one night of CLAS and/or adapted methodology.

Finally, while group-level analyses did not reveal any changes in sleep and declarative memory, there were associations between the increase in SO density during CLAS with fewer markers of sleep fragmentation (i.e., sleep fragmentation index[17], arousal). The CLAS-related SO changes were not associated with age or any other measures of sleep or memory. This may suggest that a subtype of individuals with insomnia who respond positively to CLAS may benefit from better sleep maintenance.

Future research may focus on the adaptation of CLAS methodology to individuals with insomnia and as a function of age, including the adaptation of stimulation thresholds and the use of CLAS at home and over multiple nights. In addition, a deeper investigation of responders may improve our understanding of insomnia phenotyping and the identification of subgroups of insomnia more prone to respond to CLAS. Experimental manipulation of brain oscillations using other types of sensory stimulation (e.g., rocking stimulation[21]) has been shown to improve sleep (including the generation of more SOs and spindles) and may be a suitable complementary intervention to investigate in chronic insomnia.

## Data Availability

All data produced in the present study are available upon reasonable request to the authors

## ACKNOWLEDGMENT

This research was supported by the Canadian Institutes of Health Research (awarded to Prof. Dang Vu) as well as by the Singapore National Medical Research Council (NMRC) STaR grant awarded to Prof. Michael Chee while his team was at Duke-NUS Medical School. We acknowledge the contributions of Kirsten Gong, who assisted in data preprocessing. Finally, we would like to thank the participants for giving their time and energy into this research study

## AUTHOR CONTRIBUTION STATEMENT

Conceptualization: TDV, JLO, MWLC; Data collection: JLO, TBT, ARD, NIYNC; Data curation: AAP, EMP; Formal analysis: AAP, EMP, NEC; Methodology: AP, JLO, NEC; Funding acquisition: TDV, MWLC; Writing - original draft: AAP, TDV; Writing - review & editing AAP, EMP, JLO, NEC, TBT, ARD, NIYNC, AP, MWLC, TTDV

## Notes

### Competing Interest Statement

The authors have declared no competing interest.

### Clinical Trial

ISRCTN73971514 - https://doi.org/10.1186/ISRCTN73971514

### Funding Statement

This research was funded by the Canadian Institutes of Health Research (awarded to Prof. Dang Vu) as well as by the Singapore National Medical Research Council (NMRC) STaR grant awarded to Prof. Michael Chee while his team was at Duke-NUS Medical School

### Author Declarations

Ethics Institutional Review Board of National University of Singapore (reference H-18-004) gave ethical approval for this work

## REFERENCE

Ngo H-VV, Staresina BP. Shaping overnight consolidation via slow-oscillation closed-loop targeted memory reactivation. Proc Natl Acad Sci 2022;119:e2123428119. 10.1073/pnas.2123428119.

Ngo HVV, Martinetz T, Born J, Molle M. Auditory closed-loop stimulation of the sleep slow oscillation enhances memory. Neuron 2013;78:545–53. 10.1016/j.neuron.2013.03.006.

Ong JL, Lo JC, Chee Niyn, Santostasi G, Paller KA, Zee PC, et al. Effects of phase-locked acoustic stimulation during a nap on EEG spectra and declarative memory consolidation. Sleep Med 2016;20:88–97. 10.1016/j.sleep.2015.10.016.

Diep C, Garcia-Molina G, Jasko J, Manousakis J, Ostrowski L, White D, et al. Acoustic enhancement of slow wave sleep on consecutive nights improves alertness and attention in chronically short sleepers. Sleep Med 2021;81:69–79. 10.1016/j.sleep.2021.01.044.

Ong JL, Patanaik A, Chee NIYN, Lee XK, Poh JH, Chee MWL. Auditory stimulation of sleep slow oscillations modulates subsequent memory encoding through altered hippocampal function. Sleep 2018;41. 10.1093/sleep/zsy031.

Henin S, Borges H, Shankar A, Sarac C, Melloni L, Friedman D, et al. Closed-loop acoustic stimulation enhances sleep oscillations but not memory performance. eNeuro 2019;6. 10.1523/ENEURO.0306-19.2019.

Pathak V, Juan E, Goot R van der, Talamini LM. The looping lullaby:closed-loop neurostimulation decreases sleepers’ sensitivity to environmental noise 2021:2021.08.07.455505. 10.1101/2021.08.07.455505.

Bulcke LV den, Davidoff H, Heremans E, Potts Y, Vansteelandt K, Vos MD, et al. Acoustic Stimulation to Improve Slow-Wave Sleep in Alzheimer’s Disease:A Multiple Night At-Home Intervention. Am J Geriatr Psychiatry 2025;33:73–84. 10.1016/j.jagp.2024.07.002.

Dudysová D, Janků K, Piorecký M, Hantáková V, Orendá č ová M, Piorecká V, et al. Closed-loop auditory stimulation of slow-wave sleep in chronic insomnia:a pilot study. J Sleep Res 2024;33:e14179. 10.1111/jsr.14179.

Lustenberger C, Ferster ML, Huwiler S, Brogli L, Werth E, Huber R, et al. Auditory deep sleep stimulation in older adults at home:a randomized crossover trial. Commun Med 2022;2:1–16. 10.1038/s43856-022-00096-6.

Debellemaniere E, Chambon S, Pinaud C, Thorey V, Dehaene D, Léger D, et al. Performance of an ambulatory dry-EEG device for auditory closed-loop stimulation of sleep slow oscillations in the home environment. Front Hum Neurosci 2018;12. 10.3389/fnhum.2018.00088.

American Academy of Sleep Medicine. International Classification of Sleep Disorders, Third Edition (ICSD-3). Darien, IL:American Academy of Sleep Medicine; 2014.

American Psychiatric Association. Diagnostic and Statistical Manual of Mental Disorders (DSM-V). Arlington (VA):2013. 10.1007/springerreference_179660.

Bastien CH, Vallières A, Morin CM. Validation of the insomnia severity index as an outcome measure for insomnia research. Sleep Med 2001;2:297–307. 10.1016/S1389-9457(00)00065-4.

American Academy of Sleep Medicine. The AASM Manual for the Scoring of Sleep and Associated Events:Rules, Terminology, and Technical Specifications. Version 3. Darien, IL:American Academy of Sleep Medicine; 2023.

Ray LB, Fogel SM, Smith CT, Peters KR. Validating an automated sleep spindle detection algorithm using an individualized approach. J Sleep Res 2010;19:374–8. 10.1111/j.1365-2869.2009.00802.x.

Haba-Rubio J, Ibanez V, Sforza E. An alternative measure of sleep fragmentation in clinical practice:The sleep fragmentation index. Sleep Med 2004;5:577–81. 10.1016/j.sleep.2004.06.007.

Schneider J, Lewis PA, Koester D, Born J, Ngo HVV. Susceptibility to auditory closed-loop stimulation of sleep slow oscillations changes with age. Sleep 2020;43. 10.1093/sleep/zsaa111.

Muehlroth BE, Werkle-Bergner M. Understanding the interplay of sleep and aging:Methodological challenges. Psychophysiology 2020;57:e13523. 10.1111/psyp.13523.

Marshall L, Cross N, Binder S, Dang-Vu TT. Brain Rhythms During Sleep and Memory Consolidation:Neurobiological Insights. Physiology 2020;35:4–15. 10.1152/physiol.00004.2019.

Perrault A, Khani A, Quairiaux C, Kompotis K, Franken P, Muhlethaler M, et al. Whole-Night Continuous Rocking Entrains Spontaneous Neural Oscillations with Benefits for Sleep and Memory. Curr Biol 2019;29:402-411.e3. 10.1016/j.cub.2018.12.028.

